# Long-term cooking fuel use patterns following a clean cooking intervention: Follow-up of the multi-country HAPIN trial

**DOI:** 10.64898/2026.11.17.26368324

**Authors:** Kendra N. Williams, Katherine Kearns, Stephen Mugel, Anaité Díaz-Artiga, Florien Ndagijimana, Ajay Pillarisetti, Michael A. Johnson, Amy Lovvorn, William Checkley, Kyle Steenland, Lisa M. Thompson, John P. McCracken, Ghislaine Rosa, Saritha Sendhil, Vigneswari Aravindalochanan, Kalpana Balakrishnan, Jennifer Peel, Thomas F. Clasen, the HAPIN Investigators

## Abstract

**Introduction:** The exposure and health benefits of clean cooking depend on sustained, exclusive use, yet few studies have assessed fuel use after trial support ends. We conducted a prospective follow-up of the Household Air Pollution Intervention Network (HAPIN) trial cohort to assess cooking fuel use for 4 years post-trial in India, Guatemala, and Rwanda.

**Methods:** Intervention households in the HAPIN trial received a free liquefied petroleum gas (LPG) stove, fuel, and behavioral support for 18 months. At trial completion, control participants in Guatemala and Rwanda were offered a free LPG stove or alternative compensation, and in India, a free LPG stove and 18 months of free fuel. We surveyed 2,169 participants annually for four years on exclusive and any LPG use, examined post-trial use trajectories, and assessed factors associated with post-trial LPG use using logistic regression.

**Results:** Despite near-exclusive use during the trial, post-trial, few intervention participants used LPG exclusively (29% in India, <3% in Guatemala, near 0% in Rwanda). By year 4, 82% (India), 50% (Guatemala), and 10% (Rwanda) of intervention participants used any LPG. Sustained use was highest in India; non-use was most common in Rwanda; delayed use (starting years 3–4) was most common in Guatemala. Intervention assignment (aOR 1.8 [1.3–2.7], driven by Guatemala), socioeconomic status (up to aOR 40.9 [15.7–106.6] for Q4, relative to Q1), perceived ability to use LPG (aOR 3.9 [2.6–5.7]), household size (aOR 0.86 [0.79–0.93] per member), and country (aOR 5.4 [2.3–12.5] for India; aOR 1.9 [1.3–2.7] for Guatemala, relative to Rwanda) were significantly associated with post-trial LPG use. Only 27% of Guatemalan and 7% of Rwandan control participants chose an LPG stove (or electric, in Guatemala) as compensation, citing unaffordability and inaccessibility.

**Conclusions:** A time-limited intervention, though successful in promoting LPG preference and sustained non-exclusive use in India and Guatemala, is unlikely to sustain the exclusive clean fuel use needed for long-term environmental and health benefits without policies supporting affordability and accessibility.

## Introduction

Household air pollution (HAP) remains a major public health challenge, particularly in low-and middle-income countries (LMICs).^1^ More than two billion people still rely on solid fuels such as wood, charcoal, and dung for cooking.^2^ HAP has far-reaching negative consequences, contributing to poor health outcomes, environmental degradation, lost economic and livelihood opportunities—particularly for women—and increased climate-warming emissions such as black carbon and carbon dioxide.^3,4^

The World Health Organization classifies liquefied petroleum gas (LPG) as a clean and scalable alternative to biomass for cooking.^5^ Although the evidence on health benefits is mixed,^6,7^ exclusive or near-exclusive LPG use can dramatically reduce HAP emissions and exposures, and may yield significant time savings, welfare, environmental, climate, and other benefits.^8^ Modeling indicates that near-exclusive use of LPG (i.e., using only LPG for all cooking needs) is necessary to achieve potential health benefits.^9^ However, exclusive LPG use over the long-term remains uncommon in many contexts, due in part to limited accessibility and recurring fuel costs.^10^

Rogers’ Diffusion of Innovation Theory identifies five factors driving long-term technology adoption: relative advantage (perceived benefit over existing alternatives), compatibility (fit with current practices), trialability (opportunity for low-risk experimentation), observability (visibility of results), and complexity (ease of understanding and use).^11^ Some evidence from clean cooking trials supports this framework. In Peru, a 12-month LPG intervention led to high levels of sustained, though not exclusive, LPG use in the year following the trial.^12^ In Uganda, offering a free trial period with flexible payment options increased uptake of fuel-efficient cookstoves.^13^ In Bangladesh, households were more likely to retain their LPG stove rather than exchange it for cash after a trial period.^14^ These findings suggest that free trial periods can support long-term adoption by enabling households to experience LPG’s advantages firsthand. However, rigorous, multi-year data are needed to assess long-term usage patterns and the role of household, social, market, and other contextual factors.

Our study, the Household Air Pollution Intervention Network (HAPIN) randomized controlled trial, addressed these gaps by providing an LPG stove and an 18-month supply of LPG and tracking participants for four years after the end of the trial. During the HAPIN trial, intervention participants almost completely abandoned their traditional stoves (biomass was used on 0.4% of monitored days); 96% of intervention participants reported exclusive LPG use on the prior day (n=984 of 1,029) compared to only 1% in the control group (n=9 of 1,058). Most traditional stove use in intervention households was due to running out of LPG, other household members using the traditional stove, or issues with LPG stove functionality, with very little due to cultural or social reasons.^15^ In this paper, we quantify the impact of the intervention on long-term LPG use, compare post-trial LPG use between intervention and control groups, and identify factors associated with post-trial LPG use across three diverse settings in Latin America, Africa, and Asia.

## Methods

### Study population and sites

This study examined fuel use in participants enrolled in the HAPIN study, a multi-country randomized controlled trial designed to assess the health impacts of LPG fuel use during pregnancy and early childhood. Detailed descriptions of the study design have been published elsewhere.^16,17^ In brief, the trial enrolled approximately 800 pregnant women between 9 and 19 weeks of gestation in each of four rural research sites: Jalapa, Guatemala; Puno, Peru; Kayonza, Rwanda; and Tamil Nadu, India (stratified between Villupuram and Nagapattinam districts).

Eligible participants primarily used biomass fuels for cooking at enrollment. Those randomized to the intervention group received a free LPG stove and fuel refills delivered as needed, regular behavioral support to encourage exclusive LPG use, and stove maintenance services as necessary. At the end of the trial, participants in the intervention group retained their LPG stove and one cylinder, but free LPG refills were discontinued. In India, all control group participants received an LPG stove along with 18 months of free fuel as needed after the trial concluded, although they did not receive any behavioral reinforcement for LPG use as was provided to intervention participants during the trial. In Rwanda and Guatemala, control participants were offered a choice between receiving an LPG stove and a small supply of fuel or an alternative form of compensation.^18^

### Trial sites

We conducted annual post-trial follow-up assessments in Guatemala, India, and Rwanda; Peru was not included for logistical and financial reasons. In Guatemala and Rwanda, national estimates indicate nearly universal reliance on biomass as the primary cooking fuel in rural areas (82.7% and 99%, respectively).^19^ Guatemala has made progress expanding LPG infrastructure and awareness, but access remains challenging in rural areas, and no government programs currently support LPG financing. Rwanda’s national energy strategy emphasizes LPG primarily for urban areas, resulting in limited infrastructure and availability in rural areas, and the country lacks government programs to support LPG affordability. In India, government subsidies and sustained promotion of LPG—through the ongoing Pradhan Mantri Ujjwala Yojana (PMUY), which expanded the LPG distribution network, provided digital subsidy transfers to women’s bank accounts, and conducted extensive public awareness campaigns—have contributed to substantially lower rural reliance on polluting fuels (34.3%).^19^ However, “stacking” (use of both clean and polluting fuels) is common even among those with access to affordable clean fuels; thus, primary fuel reliance does not mean exclusive use.^20^

### Follow-up data collection

Post-trial fuel use data were collected between March 2021 and May 2025. Stove and fuel use were assessed among both intervention and control participants using two survey-based methods. First, participants were asked to list all stove types used in their household over the last year and estimate the number of hours each stove was used per week. Second, participants completed a 24-hour recall survey indicating which stove(s) had been used during the previous day and for how many hours (see Table S1 for a comparison of data between these methods).

Both surveys were administered at the trial baseline visit and then repeated at annual follow-up visits after completion of the trial, when the child was 24 (year 1), 36 (year 2), 48 (year 3), and 60 (year 4) months of age; assessments of exposure and health outcomes were also conducted annually. The 24-hour recall survey was also conducted at the trial endline visit, when the child was 12 months old. At the same visit, a separate survey was administered to assess participants’ intentions for LPG use following the trial. This included questions on preferred and expected stove use, and LPG stove perceptions. Control participants’ reasons for not selecting an LPG stove as compensation were collected in Rwanda.

### Data analysis

**Participant and household characteristics** were summarized for all participants with data at one or more post-trial follow-up visits (years 1–4). Wealth status, educational attainment, and age were assessed at trial baseline; preference for and perceived ability to use LPG were from trial endline. Number of household members was averaged across follow-up years with available data; the baseline value was used if missing (n=30). Receipt of a clean cooking stove from the trial was determined using study administrative records.

**Indicators of fuel use.** We examined two indicators of fuel use: *exclusive* clean fuel use (LPG or electric stove only) and *any* clean fuel use (LPG or electric stove, alone or in combination with biomass). These indicators were compared across timepoints, groups, and countries as described below.

### LPG use among intervention participants over the post-trial follow-up period

At each annual follow-up visit (years 1–4), participants self-reported which fuel(s) they used for cooking over the prior year. Trends over time were assessed by country and compared to baseline values through numerical and visual examination of the data.

### Comparison of post-trial LPG use between intervention and control participants

Analyses differed by country based on the type of compensation control participants received at trial completion. In Guatemala and Rwanda, control participants were offered a choice between an LPG stove and starter fuel package or an alternative form of compensation, including an electric stove in Guatemala; not all participants selected the LPG stove. In these two countries, we descriptively compared exclusive and any LPG use at each post-trial visit between intervention and control participants. In India, all control participants received an LPG stove and 18 months of free fuel starting at trial end, but without the behavioral support component of the intervention. To isolate the contribution of behavioral support to initial LPG adoption in India, we compared prior-day LPG use among control participants at year 1 to prior-day use among intervention participants at trial endline. Exclusive and any LPG use were then compared between intervention (years 1–4) and control participants (years 2–4) in India.

### Proportion of cooking done with LPG by mixed-fuel users

For each post-trial annual follow-up, we identified mixed fuel users who reported using clean fuel and biomass over the prior year. For these mixed users, we calculated the proportion of self-reported total cooking time spent using LPG by dividing hours of LPG cooking by total hours of cooking across all stoves. This metric characterizes the extent to which LPG was used as a primary versus supplementary fuel among mixed users.

### LPG use trajectories

For each post-trial year, participants were first classified as LPG users if they reported using any LPG over the prior year (either exclusively or in combination with biomass). Next, participants were classified as following one of five trajectories based on their pattern of annual LPG use across years 1–4: sustained adopters (LPG use reported at all follow-up years); late abandoners (LPG use reported in first one or two years post-trial but discontinued); delayed users (no LPG use in follow-up years one and/or two, with adoption in years three or four); intermittent users (inconsistent LPG use across follow-up years); and no-post trial use (no LPG use reported at any follow-up year). Households were included if they had stove-use data in at least two of the four post-trial years. India control households were excluded in year 1 when they were receiving free LPG; they were included if they had at least two years with stove use data in years 2–4. Trajectory distributions were compared across countries and treatment arms descriptively.

### Predictors of post-trial LPG use

To identify factors associated with post-trial LPG use, we fit a multivariable logistic regression model with any LPG use across years 1-4 post-trial as the outcome (excluding year 1 for control households in India when free fuel was provided). Predictors were selected a priori based on variables hypothesized to influence fuel use behavior, drawing on existing literature and investigator expertise from each study country. Predictors included treatment assignment, socioeconomic status (SES), self-reported preference for LPG, perceived ability to use LPG, maternal age, number of household members, and country site. An SES index was developed based on factor analysis of self-reported household wealth indicators (housing materials, water and sanitation access, durable asset ownership, and maternal education), with values scaled to the range among study participants, as described previously.^21,22^ We categorized SES into quartiles, pooled across countries. In India, where most participants continued using some LPG post-trial, we fit a separate logistic regression of exclusive LPG use at year 4 on LPG accessibility, LPG affordability, SES, household size, district of residence, maternal age, and treatment assignment. Model fit was evaluated using the pseudo-R², area under the receiver operating characteristic curve (AUC), and Wald χ². Variance inflation factors were calculated for all predictors to assess multicollinearity.

## Results

### 1. Participant characteristics and fuel preferences

Of the 2,218 participants with trial endline data, 2,137 (96.3%) completed at least one post-trial fuel use survey and 2,012 (90.7%) completed the Year 4 survey. A further 32 households without a trial endline record also provided post-trial data, giving 2,169 households with any follow-up data (1,070 intervention participants and 1,099 control participants). Demographic characteristics at baseline of the HAPIN trial were similar for intervention and control participants with follow-up data (Table 1). Upon trial enrollment, none of the participants used primarily LPG or electric stoves. Households had on average 5 people (SD 1.6) and most of the participating women had primary school education or less. Socioeconomic status was highest in India (70% of households were in the highest wealth quartile) and lowest in Rwanda (68% of households were in the lowest wealth quartile). Educational attainment was highest in India (36% of women had completed secondary or higher) compared to 17% in Rwanda and 14% in Guatemala. Maternal age was slightly higher in Rwanda (27 years, SD 4.4) than in India (24 years, SD 3.8) or Guatemala (25 years, SD 4.5).

**Table 1.**
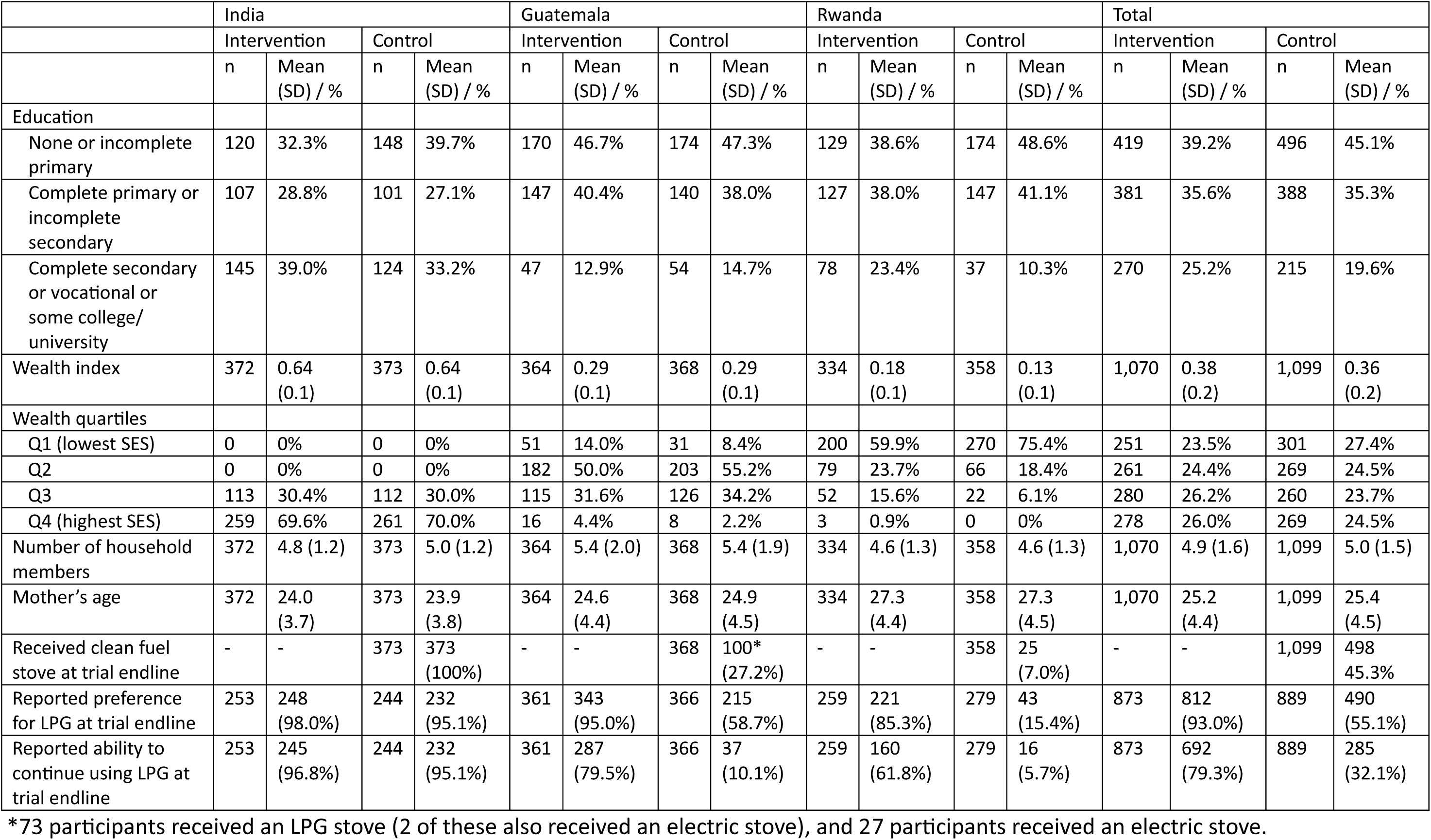
Demographic characteristics and fuel preferences of intervention and control participants by country and overall, including only participants who had data from any year of follow-up.

In India, at trial endline, >95% of both intervention and control participants said they would prefer to cook with LPG and that they would be able to continue doing so after the HAPIN trial (Table 1). Most intervention participants in Guatemala (95%) and Rwanda (85%) also said they would prefer to keep using LPG after the trial, but fewer said they would be able to continue to do so (80% in Guatemala and 62% in Rwanda). Control participants in Guatemala and Rwanda reported lower preference for LPG (59% in Guatemala and 15% in Rwanda), and much lower ability to continue using LPG (10% in Guatemala and 6% in Rwanda) than intervention participants.

Across countries, 977 of 1,762 participants (55.4%) reported planning to continue using LPG after the trial, of whom 484 (49.5%) planned exclusive use and 493 (50.5%) planned non-exclusive use. Among those planning non-exclusive use, the most cited reasons were that LPG refills are expensive (n=401, 81.3%) and that biomass is free (n=85, 17.2%), cheap (n=103, 20.9%), or easier to get (n=47, 9.5%); none cited dissatisfaction with LPG as a reason. Participants who said they did not plan to use LPG at all were not asked about their reasons.

### 2. Control group stove/fuel choices at trial endline

At the conclusion of the trial, all control participants in India received an LPG stove and 18 months of free fuel, per trial design, while in Guatemala and Rwanda, only 100 (27%) and 25 (7%) of control participants, respectively, selected an LPG stove (or electric, in Guatemala) as compensation for their trial participation (Table 1). In Rwanda, primary reasons for choosing the LPG stove were hearing good things from other HAPIN trial participants (n=9), saving money (n=7), and saving time collecting fuel and cooking (n=7). Primary reasons for not selecting LPG among control participants in Rwanda were unaffordability of LPG (n=200) and inaccessibility of LPG refills (n=71). Participants in Guatemala were not asked about their reasons for choosing LPG or not.

### 3. Post-trial LPG use among intervention participants

Following the end of the trial period, LPG use by intervention participants dropped substantially. Very few participants continued using LPG exclusively in years 1-4 after the intervention ended (Figure 1): 2-3% in Guatemala, 29% in India (43% in the first year of follow-up but only 21-27% in years 2-4), and near 0% in Rwanda (2% in the first year of follow-up and 0% thereafter).

**Figure 1.**
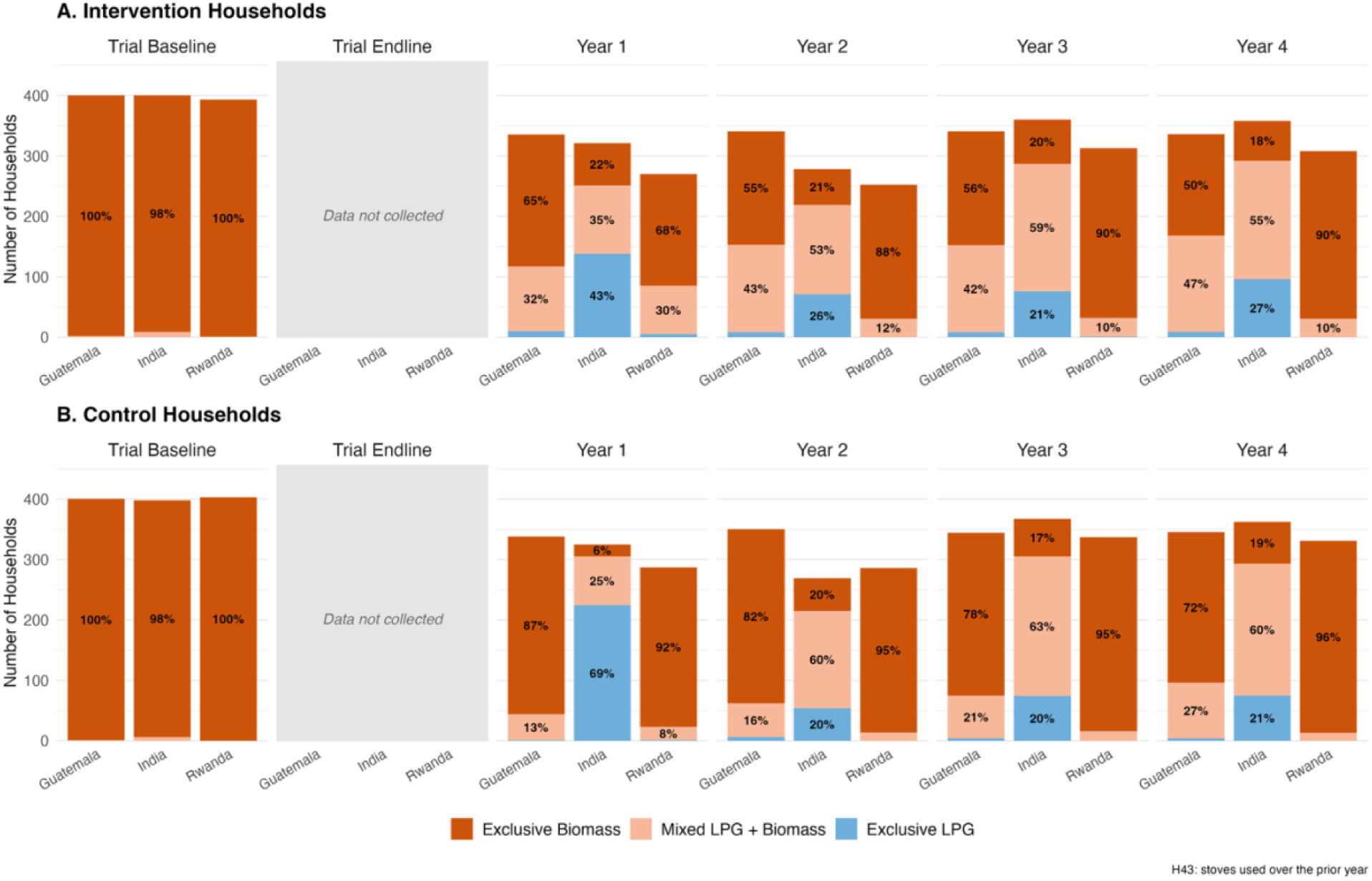
Reported stove use over the year prior to the survey among intervention and control participants, by country and timepoint. Dark orange indicates exclusive biomass stove use; light orange indicates stacking with LPG and biomass; and blue indicates exclusive LPG use. Numbers are the percentages in each category by country and round, shown only for percentages ≥5%. Data are from HAPIN trial baseline (pregnancy, 9–19 weeks’ gestation), and post-trial follow-up Years 1–4 (child ages 24, 36, 48, and 60 months, respectively).

After the trial, intervention participants had higher rates of ‘any’ LPG use (i.e., using LPG either exclusively or in combination with biomass). Reported use of any LPG in Guatemala dropped to 35% in year 1, then increased over years 2-4 post-trial to 50% in year 4. Use of any LPG decreased over time in Rwanda (from 32% in year 1 to 10% in year 4) and remained fairly stable in India (from 78% in year 1 to 82% in year 4).

### 4. Post-trial LPG use among intervention compared to control participants

Across the post-trial follow-up visits in both Guatemala and Rwanda, reported LPG use was higher among intervention participants (who received an LPG stove and 18 months of free fuel) compared to control participants (who were offered an LPG stove with limited starter fuel or other compensation at the conclusion of the trial) (Figure 1). In Guatemala, 35% of intervention and 13% of control participants reported any LPG use during the first post-trial year. In Rwanda, 32% of intervention and 8% of control participants reported any LPG use over the first year.

In Guatemala, intervention participants maintained higher use of any LPG compared to control participants after the trial, though both intervention and control groups increased LPG use over years 1-4 post-trial: by the fourth year post-trial, 50% of intervention and 28% of control participants reported any LPG use over the prior year. In contrast, in Rwanda, LPG use diminished over time. By year 4, only 10% of intervention participants and 4% of control participants reported using any LPG in the prior year. Reported exclusive LPG use remained very low across all post-trial years in both Guatemala and Rwanda for both intervention and control participants. Only 24 participants reported electric stove use post-trial; 15 did so without LPG at some follow-up visit (2 exclusively in India intervention households, and 13 in combination with biomass — 9 in Guatemala control households, 3 in India intervention households, and 1 in Guatemala intervention household). None used only electricity across all follow-up years.

In India, during year 1 post-trial (when control participants were receiving free fuel without behavioral reinforcement), control participants demonstrated similar LPG adoption as intervention participants did during the trial. During year 1 when control participants received free fuel, 85% reported exclusive LPG use and 8% reported LPG use in combination with biomass on the day prior to the survey, for a total of 93% reporting any LPG use — comparable to the 99% of intervention participants reporting any LPG use on the prior day while receiving free fuel at trial endline (Figure S1). In years 2-4 post-trial (after fuel subsidies ended), control participants in India maintained similar post-trial use of any LPG fuel as intervention participants did post-trial, with around 81% reporting any LPG use over the prior year, compared to approximately 80% of intervention participants (Figure 1). Slightly more intervention participants continued using LPG exclusively than control participants (by year 4 post-trial, 27% of intervention participants exclusively used LPG during the prior year, compared to 21% of controls).

### 5. Amount of cooking done with LPG among mixed stove users

Mixed stove users in Guatemala and India reported using LPG for slightly less than half of their cooking tasks (Figure 2). By year 4 post-trial, mixed stove users in the intervention group used LPG for 45% of cooking in India (n=196) and 38% in Guatemala (n=159); mixed stove users in the control group used LPG for 45% of cooking in India (n=218) and 40% in Guatemala (n=92). The few mixed stove users in Rwanda used LPG for a lower percentage of cooking tasks: 19% by control (n=13) and 23% by intervention (n=31) participants at year 4.

**Figure 2.**
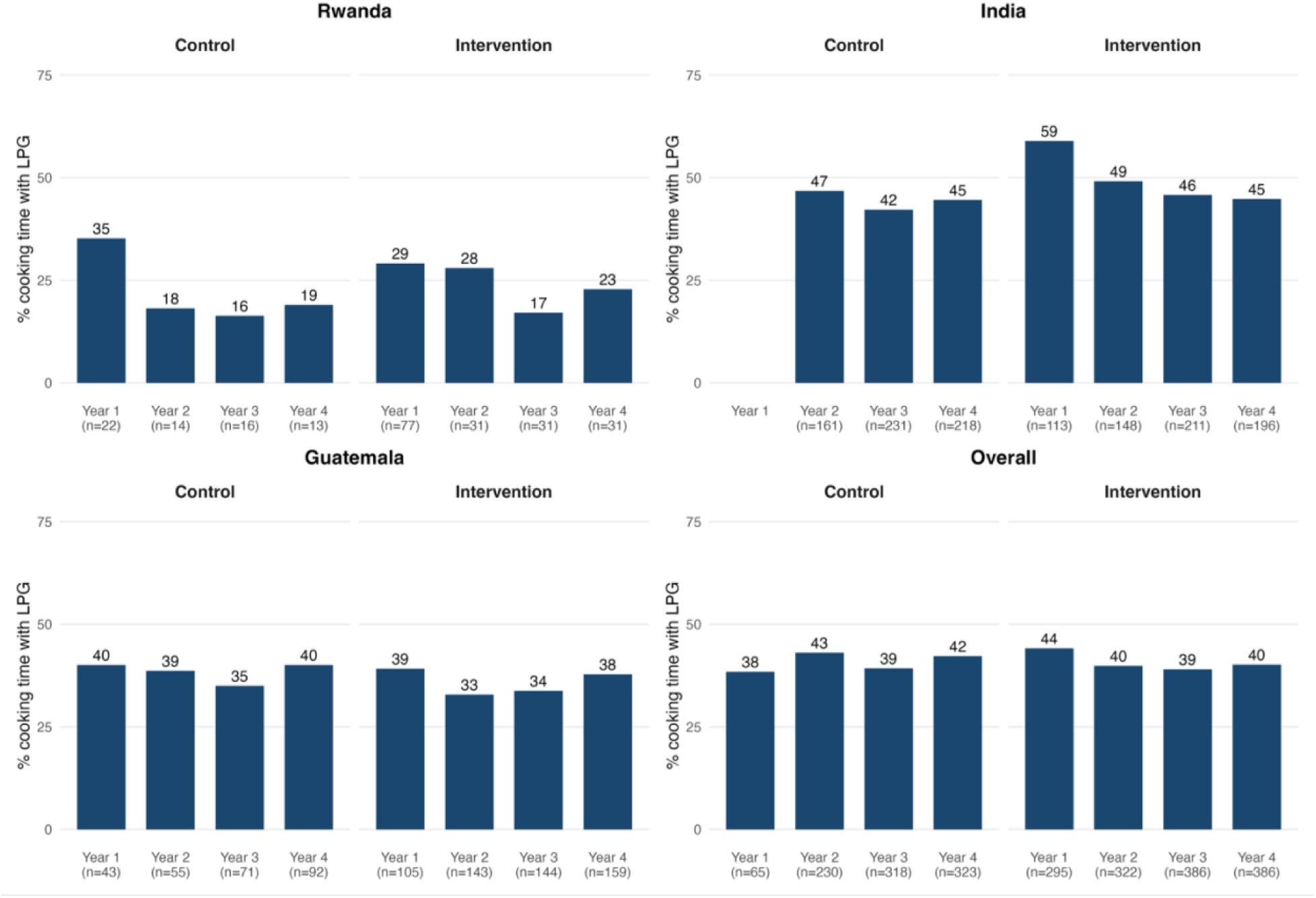
Percent of cooking done with LPG among mixed stove users, by country and treatment group. Year 1 excluded for control households in India when they were receiving free fuel.

### 6. Trajectories of LPG use

LPG use trajectories varied greatly across participants in the three countries (Figure 3; overall by country in Figure S2; n = 2,128 households with stove use data from at least two follow-up years). Rwanda had the greatest number of participants who reported no post-trial LPG use (81% across intervention and control groups) and the least number of participants who used LPG for the entire post-trial follow-up period (3%). Some intervention participants in Rwanda continued using LPG before abandoning it (17%) or continued using LPG intermittently (7%). In Guatemala, more control participants (67%) than intervention participants (34%) had no post-trial LPG use, and intervention participants had a greater proportion of sustained adopters (21% vs 8%) and intermittent LPG users (19% vs 8%). Delayed use of LPG was most common in Guatemala (17% across groups), while sustained LPG use for the entire follow-up period was highest in India (63% overall).

**Figure 3.**
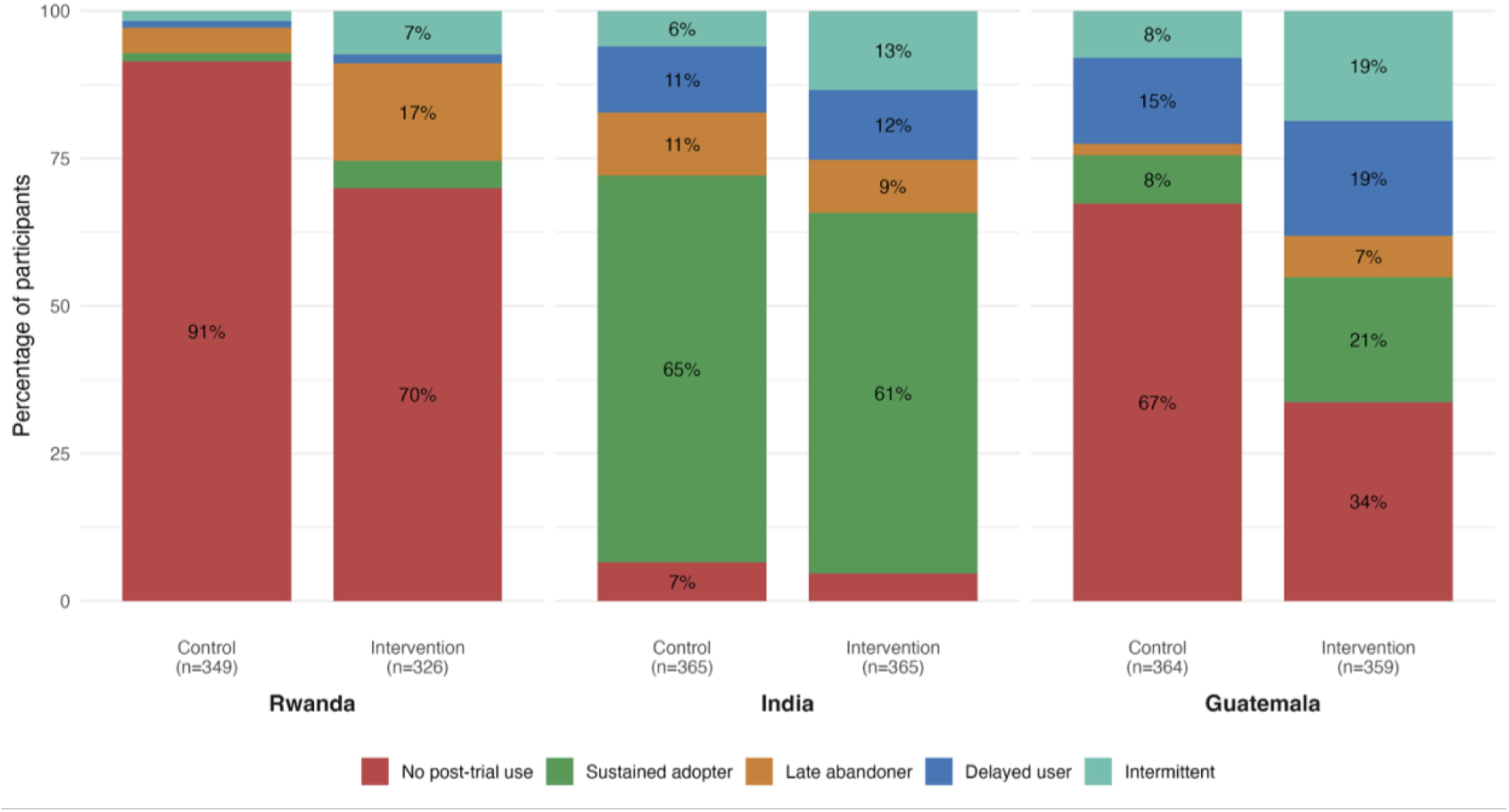
Percentage of participants in each LPG use trajectory, by country and treatment arm. Trajectories were defined based on self-reported stove use over the year prior to each survey. “No post-trial use” indicates no LPG use reported in years 1-4 (in control participants, these can be considered never-adopters); “sustained adopter” indicates LPG use reported in all follow-up years; “late abandoner” used LPG in initial post-trial years but later stopped; “delayed user” did not use LPG in initial post-trial years but began using it in later years; and “intermittent” used LPG inconsistently across follow-up years.

### 7. Factors influencing post-trial LPG use

After listwise deletion of covariates with missing values, 1,759 (81%) of the 2,169 participants with any post-trial fuel use data were included in the fully adjusted logistic regression model. The fully adjusted multivariable logistic regression model demonstrated good discrimination, with a pseudo-R² of 0.46, area under the ROC curve (AUC) of 0.91, and a Wald χ²(10) = 515.97 (p < 0.001). Variance inflation factors (VIF) for all predictors were below 5.1 (mean VIF = 2.43).

We saw a significant effect of treatment assignment, socioeconomic status, perceived ability to use LPG, number of household members, and country (Figure 4). Maternal age was not significantly associated with LPG use during the follow-up years. Assignment to the intervention arm was associated with 84% higher odds of LPG uptake compared to the control arm (adjusted OR [aOR] 1.84, 95% CI 1.26 – 2.69). Compared to the lowest SES quartile—a composite index that included maternal education along with other household assets and characteristics— participants in SES Q2 had nearly four times the odds of LPG use (aOR 3.74, 95% CI 2.47 – 5.66), those in Q3 had nearly 14 times the odds (aOR 13.73, 95% CI 8.78 – 21.47), and those in the highest SES quartile (Q4) had approximately 41 times the odds of post-trial LPG use (aOR 40.94, 95% CI 15.72 – 106.62). Participants who reported their ability to use LPG at the trial endline visit had approximately four times the odds of post-trial LPG use compared to those who did not (aOR 3.87, 95% CI 2.63 – 5.69). In contrast, stated preference for LPG at trial endline was not significantly associated with post-trial LPG use (aOR 1.26, 95% CI 0.86 – 1.84). Greater household size was significantly associated with lower odds of LPG use (aOR 0.86 per additional household member, 95% CI 0.79 – 0.93). Relative to Rwanda, participants in India had markedly higher odds of LPG use (aOR 5.38, 95% CI 2.32 – 12.51); participants in Guatemala also had higher odds compared to Rwanda (aOR 1.88, 95% CI 1.29 – 2.73).

**Figure 4.**
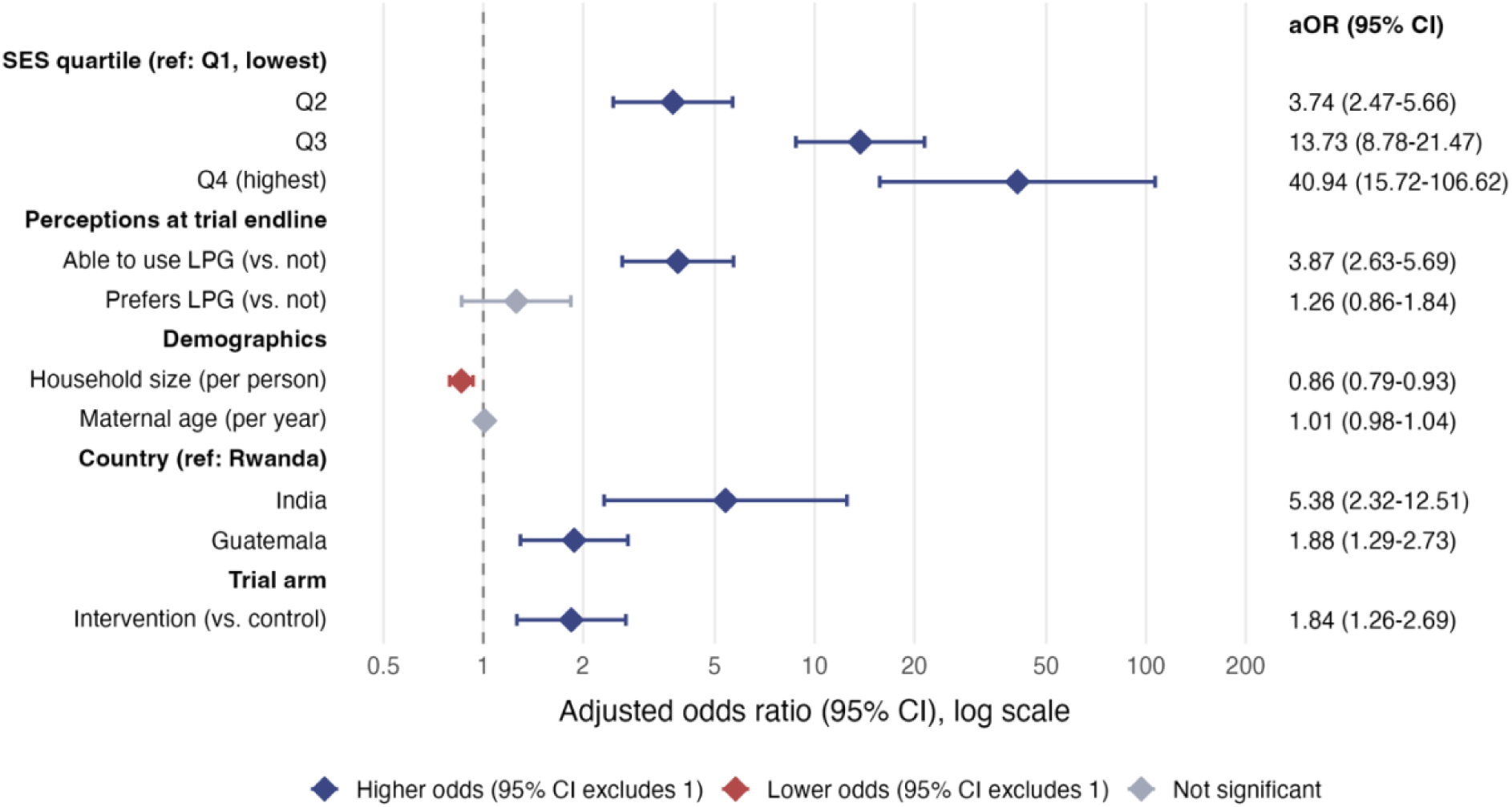
Adjusted odds ratios (aOR) and 95% confidence intervals from a multivariable logistic regression of any LPG use during post-trial follow-up (n = 1,759 households). Diamonds show aORs on a log scale and horizontal lines show 95% CIs. The values in the right-hand column are the aOR (95% CI). Blue = higher odds of LPG use, red = lower odds, grey = not significant (95% CI includes 1). SES = socioeconomic status. Reference categories: SES Q1, Rwanda, control arm. Standard errors are robust.

There were some notable differences in country-specific models (Figures S3, S4, S5). Treatment assignment was only associated with post-trial use of any LPG in Guatemala (aOR 2.08, 95% CI 1.31–3.31) (Figure S3). In both Guatemala and Rwanda, SES and perceived ability to use LPG were significant predictors; stated preference for LPG and maternal age were not. Larger household size was associated with lower odds of any LPG use in Guatemala (aOR 0.83, 95% CI 0.76–0.91) but not Rwanda (Figures S3, S4).

In India, where only 42 participants (5.7%) never used LPG post-trial, we examined predictors of exclusive LPG use at year 4 (24% of participants with year 4 data used LPG exclusively) (Figure S5). Perceived LPG accessibility and affordability were each associated with approximately twice the odds of exclusive use (aOR 2.05, 95% CI 1.34–3.13 and aOR 2.09, 95% CI 1.35–3.24, respectively). SES was not independently associated. Larger households (aOR 0.66, 95% CI 0.55– 0.80), residence in Villupuram versus Nagapattinam (aOR 0.34, 95% CI 0.23–0.52), and older maternal age (aOR 0.95, 95% CI 0.90–1.00) were each associated with lower odds of exclusive use. Post-trial exclusive LPG use was too rare in Guatemala and Rwanda to model.

## Discussion

While almost all intervention participants used LPG fuel exclusively during the HAPIN trial when fuel was free and delivered directly to the home, exclusive use was limited after the trial, when households had to pay for and procure fuel. By year 4 post-trial, only about a quarter of participants in India were still using LPG exclusively and almost none in Guatemala or Rwanda were. The trial did, however, result in substantial non-exclusive use of LPG post-trial in India and Guatemala, but not in Rwanda. LPG use trends in the four years post-trial varied by site: LPG use increased in Guatemala, remained stable in India, and declined in Rwanda. In India, behavioral reinforcement may have led to slightly higher exclusive use of LPG post-trial among intervention participants compared to control, but similar levels of non-exclusive use.

Differences in post-trial LPG use across countries were likely shaped by broader country contexts, including infrastructure, policies and political support, subsidization, average per capita income, and social norms. In India, government subsidies and active promotion of LPG created both financial support and social pressure to use it, likely contributing to higher sustained use. The PMUY program in India expanded LPG access through a network of cylinder-refill distributors, digital delivery of subsidy credits to women’s bank accounts, and public awareness campaigns promoting LPG use; however, the scope and level of subsidy support fluctuated over the study period and varied across states. In contrast, Rwanda’s low LPG use likely reflects the absence of government programs to support LPG affordability, and a national energy strategy that emphasizes LPG for urban areas only, resulting in limited LPG infrastructure and availability outside major cities. In Guatemala, LPG infrastructure is expanding, reflected in a growing presence of LPG companies and advertising in rural areas. These trends may be driving greater awareness of and interest in adopting LPG. Nevertheless, access remains challenging in many rural areas, and no government programs currently support LPG financing in Guatemala. Differences in SES across sites also likely influenced post-trial LPG use, with higher SES in India enabling continued purchase of subsidized LPG, low SES in Rwanda limiting purchasing power, and intermediate SES in Guatemala contributing to moderate continued LPG use.

Other trials have similarly found near-exclusive use of LPG during the intervention when it was provided and delivered for free, with lower post-trial use that was higher in areas with LPG subsidies. In Ghana, where LPG fuel subsidies were not available, the GRAPHS study found that no households reported LPG stove use three years after an 18-month LPG trial in which free fuel was delivered.^23^ The CHAP study in Peru, where LPG subsidies were available, found that intervention participants continued using LPG on 85% of monitored days but only exclusively for 51% of days after a 12-month LPG trial with free LPG fuel and deliveries.^12^ Other studies have found that pay as you go schemes with lower, flexible incremental prices also increase LPG use.^24^ These studies suggest that cost, not social or cultural preferences, is the primary driver of LPG use, and that efforts to subsidize LPG and create a more accessible payment structure are most likely to facilitate its adoption.

At an individual level, factors influencing post-trial LPG use align with other literature demonstrating higher use in households with greater socioeconomic status and maternal education, and lower use in larger households.^10^ Although the majority of intervention participants preferred to continue using LPG after the trial ended, fewer said they would be able to do so, primarily due to the ongoing cost of fuel refills. Our regression results also showed that lower SES and larger household size were associated with lower post-trial LPG use. Other studies have similarly found affordability to be a main driver of sustained clean fuel use.^25^ Notably, the HAPIN, GRAPHS and CHAP trials found no protective effects across a range of primary health outcomes, making it unclear whether real or perceived health gains motivated continued use in any setting. Other work has found low post-trial adoption of interventions with demonstrated health benefits that require significant resources, as well as high post-trial adoption of “ineffective” interventions with null health benefits, suggesting health motivation alone is insufficient to sustain behavior change in the absence of structural support.^26^

## Strengths and Limitations

This study has several strengths. Longitudinal follow-up allowed assessment of the longer-term effects of an 18-month free LPG fuel, home delivery, and behavioral reinforcement intervention on fuel use patterns. The multi-country design (Guatemala, India, Rwanda) captures diverse sociocultural, economic, and infrastructural contexts. High retention (96% of participants with trial endline data completed at least one post-trial fuel use survey and 91% completed the year 4 survey) and repeated measures enable analysis of trends and trajectories over time.

Participants’ long-standing trust in field staff increased confidence in self-reported data. These findings have clear policy relevance, demonstrating that even short-term provision of free LPG can substantially increase long-term LPG use where infrastructure is sufficient, although exclusive use remains limited.

Several limitations should also be noted. First, post-trial fuel use was measured annually at the household level, which limits the ability to capture within-household seasonal variation; however, because surveys were administered across different households throughout the year, the pooled data capture fuel use across all seasons. Second, all data were self-reported, introducing potential recall and social desirability bias. Participants may have overreported LPG use to align with perceived study expectations or underreported LPG use to emphasize financial need in hopes of receiving further benefits from the project. Ongoing contact through health and exposure monitoring may have heightened this bias, or improved recall. Although retention was high, participants lost to follow-up may differ systematically from those who remained in the study. Control participants may not represent the general population, as their prior trial participation likely increased awareness of LPG benefits. Third, in Guatemala and Rwanda, few control participants selected an LPG stove as compensation for trial completion. Thus, our analyses in these countries compare the LPG intervention to the option of receiving an LPG stove rather than to actual stove provision; this low uptake likely reflects limited prioritization of LPG and suggests that sustained use may have remained low even with stove distribution. Those who elected to receive the LPG stove may be different from those who chose another compensation option. Fourth, we did not collect qualitative data to contextualize our results or confirm our interpretation of the data. Finally, since HAPIN trial participants received free fuel delivered to their homes during the trial, they may have developed an expectation that they should not have to pay or arrange for the delivery of LPG post-trial, influencing their fuel usage behaviors.

## Conclusion

The 18-month HAPIN trial was successful in achieving awareness and stated preference for cooking with LPG, as well as continued use of LPG in combination with biomass in India and Guatemala over four years post-trial. However, exclusive use of LPG four years after the trial was limited to about one quarter of participants in India, and very few participants in Rwanda continued using any LPG. Fuel access and affordability were key barriers to long-term adoption. Additional efforts beyond a short subsidization and behavioral support period provided during the trial, such as national-level policies that enhance LPG accessibility and subsidize fuel costs, are likely needed to achieve greater sustained and exclusive LPG use.

## Funding

Research reported in this publication was supported by the National Heart, Lung and Blood Institute under Award Number UM1HL134590 in collaboration with the Bill & Melinda Gates Foundation [OPP1131279], and by the National Institute of Environmental Health Sciences under Award Numbers R56ES033530 and R01ES033530, of the National Institutes of Health. The total cost of the project described in this manuscript was approximately $37M, all of which was administered as federal funds. It is subject to the NIH Public Access Policy. Through acceptance of this federal funding, NIH has been given a right to make this manuscript publicly available in PubMed Central upon the Official Date of Publication, as defined by NIH. Under the grant conditions of the Gates Foundation, a Creative Commons Attribution 4.0 License has already been assigned to the Author Accepted Manuscript version that might arise from this submission. The content is solely the responsibility of the authors and does not necessarily represent the official views of the National Institutes of Health or the Gates Foundation.

## Ethical Approval

The study protocol was reviewed and approved by institutional review boards (IRBs) or Ethics Committees at Emory University (00089799), Johns Hopkins University (00007403), Sri Ramachandra Institute of Higher Education and Research (Deemed University) (IEC-NI/16/JUL/54/49; IEC-NI/21/OCT/80/120) and the Indian Council of Medical Research–Health Ministry Screening Committee (5/8/4-30/(Env)/Indo-US/2016-NCD-I; 2021-8184), Universidad del Valle de Guatemala (146-08-2016) and the Guatemalan Ministry of Health National Ethics Committee (11-2016), the London School of Hygiene and Tropical Medicine (11664; 22970) and the Rwandan National Ethics Committee (853/RNEC/2016; 317/2017; 357/2018; 194/2019; 929/2020; 64/2021; 45/2022; 47/2023; 17/2024; 666/2024), and Washington University in St. Louis (201611159). Written informed consent was obtained from all participants. The HAPIN trial is registered with ClinicalTrials.gov (Identifier NCT02944682).

## Data Availability

All data produced in the present study are available upon reasonable request to the authors

## Acknowledgments

The investigators would like to thank the members of the advisory committee – Drs. Patrick Breysse, Donna Spiegelman, and Joel Kaufman - for their valuable insight and guidance throughout the implementation of the trial. We also wish to acknowledge all research staff and study participants for their dedication to, and participation in, this important trial.

A multidisciplinary, independent Data and Safety Monitoring Board (DSMB) appointed by the NIH National Heart, Lung, and Blood Institute (NHLBI) monitored the quality of the data and protected the safety of participants enrolled in the HAPIN trial. The DSMB consisted of: Catherine Karr (Chair), Nancy R. Cook, Stephen Hecht, Joseph Millum, Nalini Sathiakumar (deceased), Paul K. Whelton, and Gail Weinmann and Thomas Croxton (Executive Secretaries). Program Coordination: Gail Rodgers, Bill & Melinda Gates Foundation; Claudia L. Thompson, National Institute of Environmental Health Sciences; Mark J. Parascandola, National Cancer Institute; Marion Koso-Thomas, Eunice Kennedy Shriver National Institute of Child Health and Human Development; Joshua P. Rosenthal, Fogarty International Center; Concepcion R. Nierras, NIH Office of Strategic Coordination–The Common Fund; Katherine Kavounis, Dong-Yun Kim, Barry S. Schmetter (deceased), and Antonello Punturieri, NHLBI. This research represents the NIH’s contribution to the Global Alliance for Chronic Diseases (GACD) coordinated call for research on prevention and management of chronic lung diseases for 2016.

## Notes

### Competing Interest Statement

The authors have declared no competing interest.

### Clinical Trial

NCT02944682

### Author Declarations

The study protocol was reviewed and approved by institutional review boards (IRBs) or Ethics Committees at Emory University (00089799), Johns Hopkins University (00007403), Sri Ramachandra Institute of Higher Education and Research (Deemed University) (IEC-NI/16/JUL/54/49; IEC-NI/21/OCT/80/120) and the Indian Council of Medical Research-Health Ministry Screening Committee (5/8/4-30/(Env)/Indo-US/2016-NCD-I; 2021-8184), Universidad del Valle de Guatemala (146-08-2016) and the Guatemalan Ministry of Health National Ethics Committee (11-2016), the London School of Hygiene and Tropical Medicine (11664; 22970) and the Rwandan National Ethics Committee (853/RNEC/2016; 317/2017; 357/2018; 194/2019; 929/2020; 64/2021; 45/2022; 47/2023; 17/2024; 666/2024), and Washington University in St. Louis (201611159).

