## Supplemental Materials for "Long-term cooking fuel use patterns following a clean cooking intervention: Follow-up of the multi-country HAPIN trial"

### Supporting Information

Figure S1. Reported fuel use on the day prior to the survey among intervention and control participants, by country and timepoint. At trial endline, intervention participants were still receiving free fuel. At trial endline, participants were only asked about stoves used on the day prior to the survey; they were not asked about stoves used overall during the previous year. This data is included for comparisons to trial endline stove use.

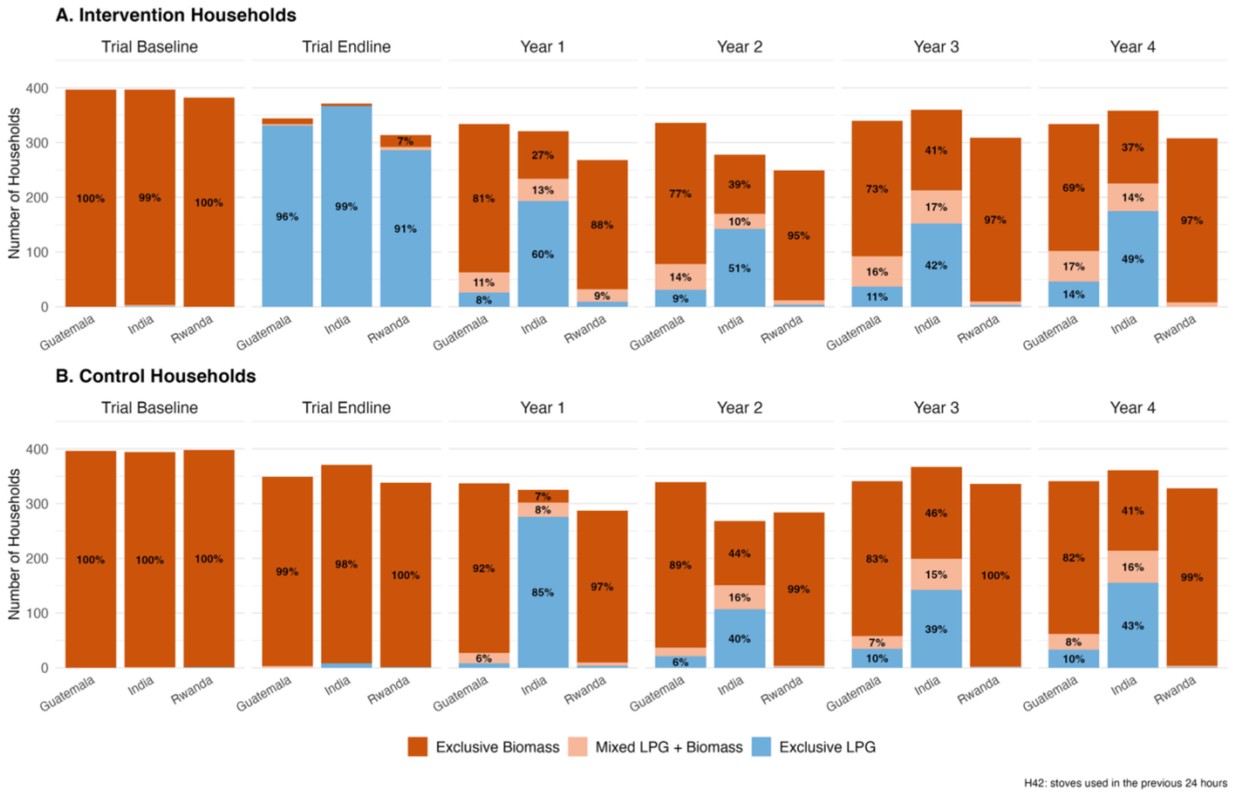

Figure S2. Percentage of participants in each LPG use trajectory, by country. Trajectories were defined based on self-reported stove use over the year prior to each survey. “No post-trial use” indicates no LPG use reported in years 1-4 (in control participants, these can be considered never-adopters); “sustained adopter” indicates LPG use reported in all follow-up years; “late abandoner” used LPG in initial post-trial years but later stopped; “delayed user” did not use LPG in initial post-trial years but began using it in later years; and “intermittent” used LPG inconsistently across follow-up years.

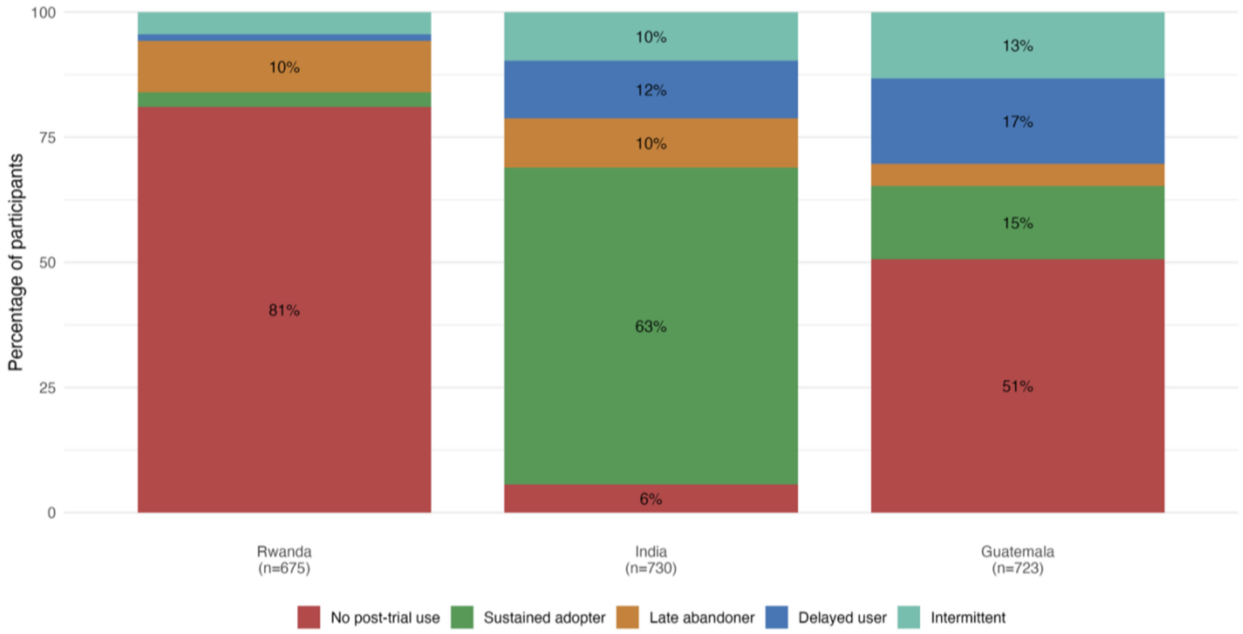

Figure S3. Predictors of ever using LPG in any of the four years post-trial, Guatemala.

Adjusted odds ratios (aOR) and 95% confidence intervals from a multivariable logistic regression of any LPG use during post-trial follow-up among households in Guatemala (n = 727). Diamonds show aORs on a log scale, horizontal lines show 95% CIs, and the right-hand column gives the aOR (95% CI). Blue indicates higher odds of LPG use, red lower odds, and grey no significant association (95% CI includes 1); the dashed line marks OR = 1. Reference categories: control arm, SES quartile Q1 (lowest). SES = socioeconomic status. Robust standard errors. Model fit: AUC = 0.79; pseudo-$R^2$ = 0.20; Wald $\chi^2(8)$ = 154.14 ($p < 0.001$). Variance inflation factors (VIF) for all predictors were below 2.8 (mean VIF = 1.79).

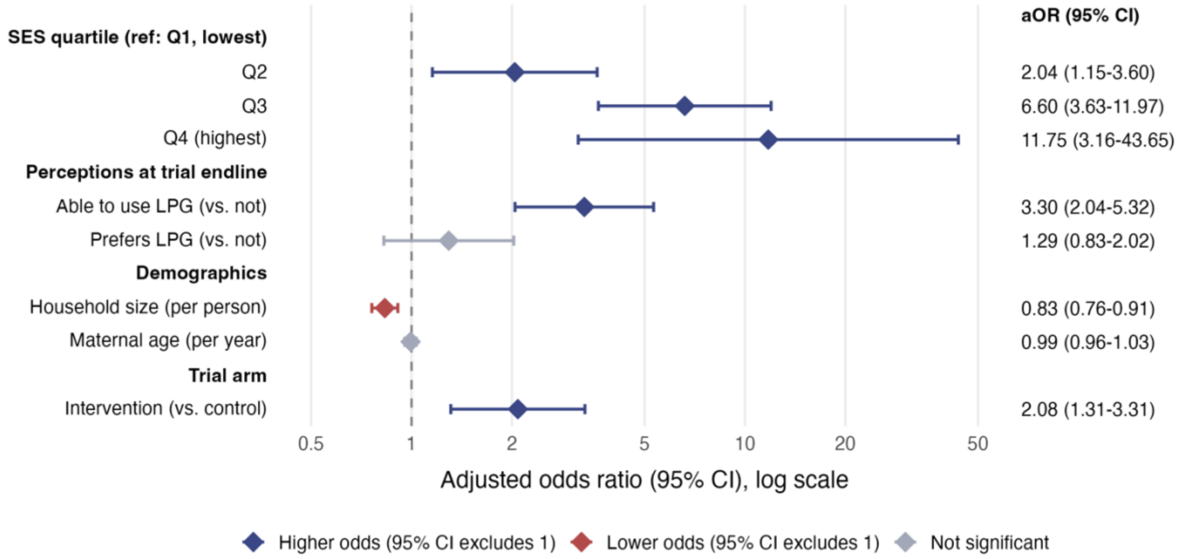

Figure S4. Predictors of ever using LPG in any of the four years post-trial, Rwanda.

Adjusted odds ratios (aOR) and 95% confidence intervals from a multivariable logistic regression of any LPG use during post-trial follow-up among households in Rwanda (n = 536). Diamonds show aORs on a log scale, horizontal lines show 95% CIs, and the right-hand column gives the aOR (95% CI). Blue indicates higher odds of LPG use, red lower odds, and grey no significant association (95% CI includes 1); the dashed line marks OR = 1. Reference categories: control arm, SES quartile Q1 (lowest). SES Q4 (highest) was omitted from the model (2 observations) because it perfectly predicted the outcome. Robust standard errors. Model fit: AUC = 0.87; pseudo-$R^2$ = 0.35; Wald $\chi^2(7)$ = 125.89 ($p < 0.001$). VIF for all predictors were below 2.3 (mean VIF = 1.53).

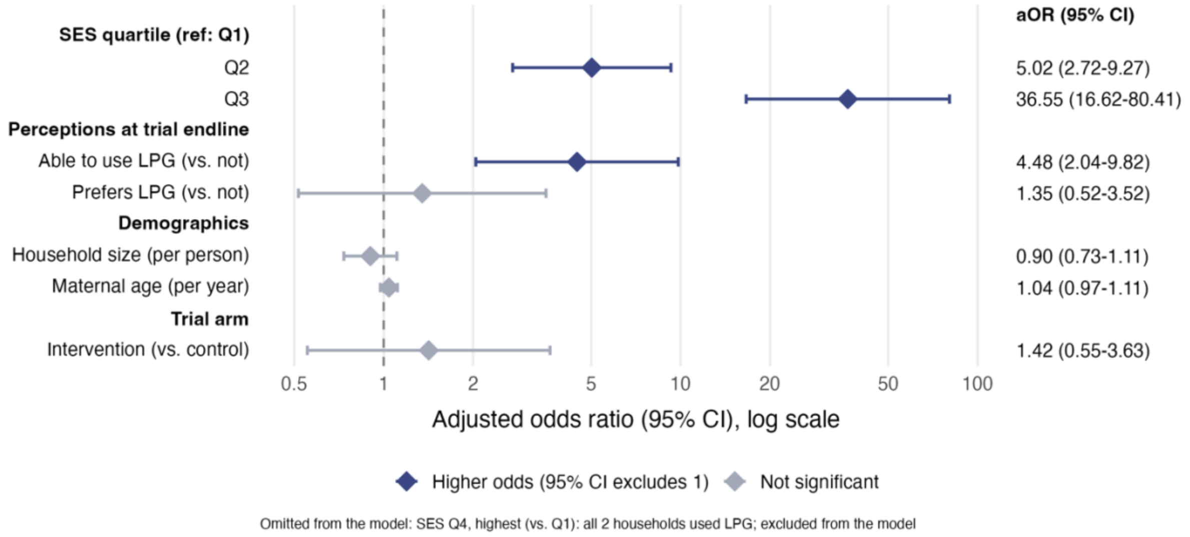

Figure S5. Predictors of exclusive LPG use in year 4 post-trial, India.

Adjusted odds ratios (aOR) and 95% confidence intervals from a multivariable logistic regression of exclusive LPG use in year 4 among households in India (n = 682). Exclusive use was the outcome because only 42 of 741 participants (5.7%) never used LPG during post-trial follow-up. Diamonds show aORs on a log scale, horizontal lines show 95% CIs, and the right-hand column gives the aOR (95% CI). Blue indicates higher odds of exclusive LPG use, red lower odds, and grey no significant association (95% CI includes 1); the dashed line marks OR = 1. Reference categories: control arm, SES quartile Q3 (no participants in India were in Q1 or Q2), Nagapattinam. Robust standard errors. Model fit: AUC = 0.72; pseudo-$R^2$ = 0.11; Wald $\chi^2(7)$ = 70.39 ($p < 0.001$). VIF for all predictors were below 1.3 (mean VIF = 1.07).

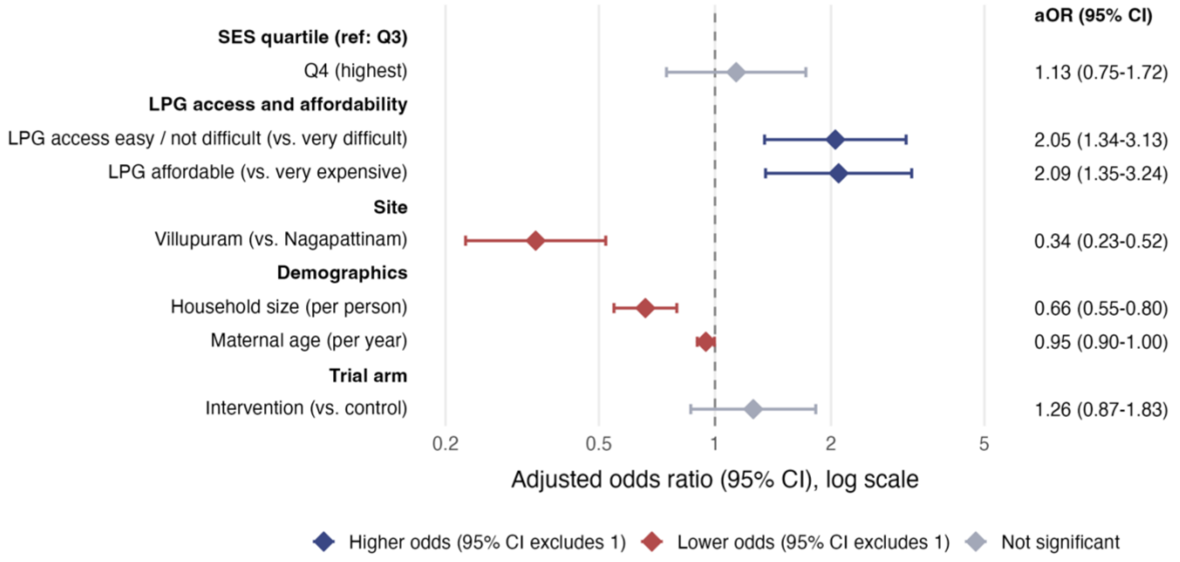

Table S1. Agreement between reported LPG use over the prior year and on the prior day, post-trial Years 1–4.

Reports of stove use over the prior year were largely consistent with reports of use on the prior day. Across 7,708 participant-visits in post-trial Years 1–4 (2,168 participants), agreement was 86.7% (Cohen's κ = 0.72, 95% CI 0.70–0.73), with κ ranging from 0.67 to 0.81 across years. Disagreement was asymmetric: in 30.4% of visits where LPG use over the prior year was reported, no LPG was used on the prior day, indicating that prior-day reports under-represent intermittent LPG use. Only about 0.5% of participants reported no LPG stove use in the past year but did report LPG use the previous day. We therefore rely primarily on prior-year reports, except for comparisons with trial endline, when only prior-day data were collected.

| | **Visits (n)** | **Participants (n)** | **Agreement** | **κ (95% CI)** | **Visits with prior-year LPG use but none on prior day** | **Visits with prior-day LPG use but none in prior year** |
|---|---|---|---|---|---|---|
| Year 1 | 1,872 | 1,872 | 90.8% | 0.81 (0.78–0.83) | 19.9% | 0.4% |
| Year 2 | 1,754 | 1,754 | 85.8% | 0.68 (0.64–0.72) | 35.2% | 0.4% |
| Year 3 | 2,053 | 2,053 | 84.7% | 0.67 (0.64–0.70) | 35.1% | 0.5% |
| Year 4 | 2,029 | 2,029 | 85.6% | 0.70 (0.67–0.73) | 31.9% | 0.5% |
| **Overall** | **7,708** | **2,168** | **86.7%** | **0.72 (0.70–0.73)** | **30.4%** | **0.5%** |

LPG use = any LPG stove used, alone or with biomass. κ = Cohen's kappa. CIs are from 2,000 bootstrap resamples of participants. The "Overall" row pools Years 1–4 (participants contribute up to four visits). 2,168 of the 2,169 participants with any post-trial fuel-use data had at least one visit with both measures. Visits with missing or unclassified data on either measure were excluded.
